# Modeling the potential impacts of outpatient antiviral treatment in reducing influenza-associated hospitalizations in the United States

**DOI:** 10.1101/2024.10.17.24315294

**Authors:** Sinead E. Morris, Sarabeth M. Mathis, Emily Reeves, Jessie R. Chung, Rebecca K. Borchering, Nathaniel M. Lewis, Svetlana Masalovich, Shikha Garg, Timothy M. Uyeki, A. Danielle Iuliano, Mark W. Tenforde, Carrie Reed, Matthew Biggerstaff

## Abstract

**Background:** Seasonal influenza causes an estimated 100,000–710,000 hospitalizations annually in the United States (U.S.). Treatment with antiviral medications, such as oseltamivir, can reduce risks of hospitalization among people with influenza-associated illness. The U.S. Centers for Disease Control and Prevention recommends initiating antiviral treatment as soon as possible for outpatients with suspected or confirmed influenza who have severe or progressive illness or are at higher risk of influenza complications.

**Methods:** We developed a probabilistic model to estimate the impact of antiviral treatment in reducing hospitalizations among U.S. outpatients with influenza. Parameters were informed by seasonal influenza surveillance platforms and stratified by age group and whether individuals had a medical condition associated with higher risk of influenza complications. We modeled different scenarios for influenza antiviral effectiveness and outpatient testing and prescribing practices, then compared our results to a baseline scenario in which antivirals were not used.

**Results:** Across the modeled scenarios, antiviral treatment resulted in 1,215–14,184 fewer influenza-associated hospitalizations on average compared to the baseline scenario (a 0.2%–2.7% reduction). The greatest effects occurred among adults ≥65 years and individuals with conditions associated with higher risk of influenza complications. Modeling 50% improvements in access to care, testing, prescribing, and treatment resulted in greater potential impacts, with over 71,000 (13.3%) influenza-associated hospitalizations averted on average compared to baseline.

**Conclusions:** Our results support recommendations to prioritize outpatient antiviral treatment among older adults and others at higher risk of influenza complications. Improving access to prompt testing and treatment among outpatients with suspected influenza could reduce hospitalizations substantially.

**Summary:** We probabilistically modeled influenza antiviral treatment among outpatients with influenza virus infection in the United States. Antiviral treatment reduced influenza-associated hospitalizations, with the greatest impacts among adults ≥65 years and persons with conditions associated with higher risk of influenza complications.

## Introduction

Seasonal influenza causes substantial morbidity in the United States, with an estimated 100,000– 710,000 influenza-associated hospitalizations occurring each year^1^. Antivirals, including neuraminidase inhibitors like oseltamivir, can reduce the risk of hospitalization for those who are ill with influenza, particularly when administered within 48 hours of symptom onset^2–4^. The U.S. Centers for Disease Control and Prevention (CDC) recommends starting antiviral treatment as soon as possible for any outpatient with suspected or confirmed influenza who is at higher risk of influenza complications or has severe or progressive illness^5^. Those considered at higher risk of influenza complications include children <2 years, adults ≥65 years, pregnant people, and individuals with chronic medical conditions including asthma, lung disease, and heart disease^6^.

Although antiviral prescribing for people with suspected or confirmed influenza is generally high among hospitalized patients, it can be low in outpatient settings, even among patients at higher risk of complications^7–9^. Antiviral prescribing also depends on testing practices and individual access to care. For example, patients seeking care within 48 hours of symptom onset may be more likely to receive an antiviral prescription than patients seeking care after 48 hours^10,11^. Similarly, patients with a positive influenza test result may be more likely to receive a prescription than patients with a negative result, or patients who are not tested^7,10^. It is unclear how these different factors influence the impact of antivirals in reducing influenza-associated hospitalizations among outpatients.

We developed a probabilistic model to estimate the impact of influenza antiviral treatment in reducing hospitalizations among outpatients with symptomatic influenza virus infection. The model is stratified by age and risk of influenza complications. We used data from seasonal influenza surveillance platforms to inform parameter inputs and modeled a range of scenarios that reflected plausible variations in antiviral effectiveness, and current testing and prescribing practices. We also explored how the impact of influenza antivirals may change with improvements in individual access to care and clinical testing and prescribing rates. Our results can be used to identify groups that may benefit most from influenza antiviral treatment.

## Methods

### Model framework

We developed a probabilistic model to track care-seeking, testing, antiviral prescribing, and risks of hospitalization among individuals with symptomatic influenza virus infection in the United States (Figure 1). The model was stratified into five age groups (0–4, 5–17, 18–49, 50–64, and ≥65 years). Within each age group, individuals were further stratified by risk of hospitalization based on the presence or absence of a condition associated with increased risk of influenza-associated complications^5^. Within each age group and risk stratification, individuals with symptomatic infection were partitioned into those who did or did not seek care in an outpatient clinical setting. For those who did not seek care, the probabilities of hospitalization were *p*_*LR*_ and *p*_*HR*_ for the low- and high-risk stratifications, respectively. Those that did seek care were partitioned according to whether they sought care within 48 hours of symptom onset (referred to as ‘early care-seekers’) or more than 48 hours after symptom onset (‘late care-seekers’).

**Figure 1.**
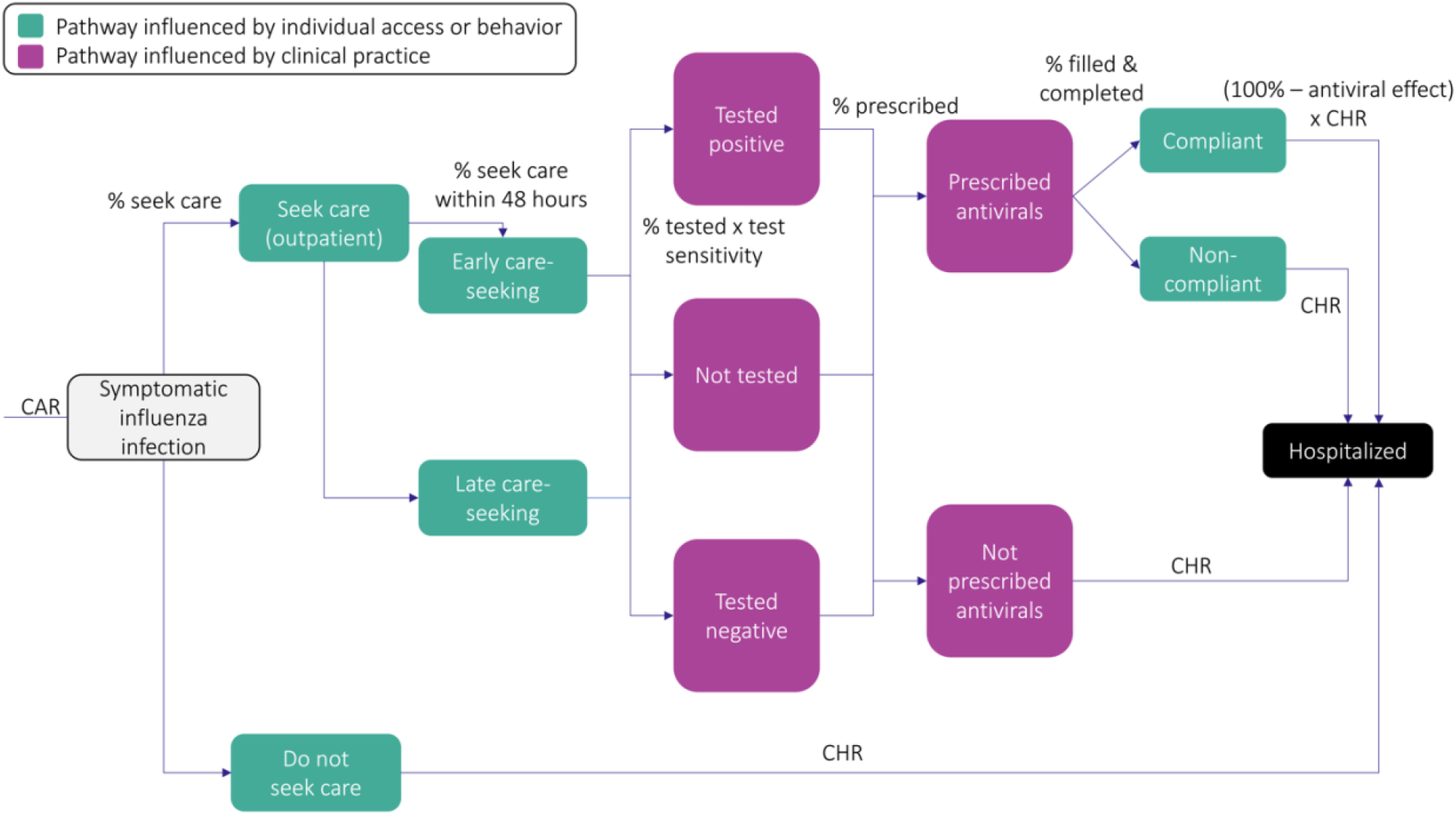
Model structure within each age group and risk stratification. The risk of hospitalization for individuals with symptomatic influenza infection depends on whether individuals seek care, the timing of care-seeking, the occurrence and result of testing, the prescribing of antiviral treatment, and the filling and completion of any treatment course. The number of individuals progressing through each pathway may change by age and/or risk stratification (see Figure S7 for a Sankey diagram showing example flows for high-risk adults ≥65 years^13^). In addition, the number progressing through each testing and prescribing pathway will vary by the timing of care-seeking (i.e., early vs. late). Similarly, antiviral effectiveness varies by the timing of care-seeking and will lead to different numbers progressing through the fully compliant to hospitalized pathway. Finally, the colors indicate compartments that are influenced by parameters relating to individual behavior and/or access to medical care and treatment (care-seeking, timing of care-seeking, and antiviral compliance) or parameters relating to clinical testing and prescribing practices. We investigate the impact of changing these parameters as part of our analyses. Abbreviations: CAR = clinical attack rate; CHR = case-hospitalization ratio.

Both early and late care-seekers had a certain probability of being tested for influenza in the outpatient setting and, if tested, a certain probability of testing positive that was determined by the sensitivity of the diagnostic assay. Early and late care-seekers were subsequently partitioned into three groups: those who tested positive for influenza, those who tested negative, and those who did not receive a test. We assumed individuals in these groups were then prescribed antivirals with probabilities determined by their test status, timing of care-seeking, and whether they were in a low- or high-risk stratification. Those who were not prescribed antivirals were hospitalized with probabilities *p*_*LR*_ and *p*_*HR*_ for the low- and high-risk stratifications, respectively. Those who were prescribed antivirals but did not fill their prescription or did not complete the full treatment course were also hospitalized with probabilities *p*_*LR*_ and *p*_*HR*_. Those who did fill their prescription and complete the full course (referred to as ‘full antiviral compliance’) were hospitalized with reduced probabilities (1 − *r*) *p*_*LR*_ and (1 − *r*) *p*_*HR*_, where *r* is the effectiveness of antivirals in reducing the risk of influenza-associated hospitalization. We parameterized *r* based on reported effectiveness of oseltamivir, the most commonly prescribed influenza antiviral in outpatient settings^12^, and allowed it to vary with the timing of care-seeking^3,4^.

### Parameterization

We determined the number of individuals in each age group and risk stratification using 2022 US census data and age-specific estimates of the proportion of people at higher risk of influenza complications^14,15^ (Table S1). The latter estimates included pregnant people and those with chronic medical conditions including asthma, heart disease, and lung disease, but did not include people with a body mass index (BMI) ≥40 or those living in long-term care facilities who are also considered to be at higher risk of influenza complications^5^. The number of individuals developing symptomatic influenza virus infection was approximated using age-specific clinical attack rates for influenza-associated illness during the 2022– 2023 influenza season^16^. Age-specific risks of hospitalization were obtained from influenza-associated case-hospitalization ratios and adjusted for risk stratification^17–19^ (Table S1). Probabilities of seeking care, being an early or late care-seeker, and filling and completing an antiviral prescription were informed by seasonal influenza surveillance platforms and prior literature^7,11,20–23^. Care-seeking varied by age and risk stratification, the timing of care-seeking varied by age but was constant across risk stratifications, and antiviral compliance was constant across age and risk stratifications^11^.

The remaining parameters of testing, prescribing, and antiviral effectiveness are highly variable in outpatient settings. We therefore defined a range of scenarios that reflected plausible values for these parameters and were informed by prior literature and unpublished data from seasonal influenza surveillance platforms (Table 1). We defined three scenarios (‘low’, ‘intermediate’, and ‘high’) for the probability an outpatient with symptomatic influenza virus infection was tested for influenza^21,22^. For each scenario, we assumed these probabilities were the same for all age groups and risk stratifications but were higher among early care-seekers compared with late care-seekers. To determine the proportion of outpatients testing positive for influenza, we multiplied the testing probability by estimates of test sensitivity for rapid molecular assays^24,25^. We also defined two scenarios (‘low’ and ‘high’) for the probability an outpatient was prescribed influenza antiviral medications. We assumed these probabilities were constant across age groups, but varied with risk stratification, time of care-seeking, and the result of influenza testing^7,10,21,22,26^. Finally, we explored three scenarios for antiviral effectiveness (‘low’, ‘intermediate’, and ‘high’) that were primarily informed by studies of oseltamivir but could be applicable to other existing or novel influenza antivirals given the broad range of values considered^2–4,27–29^. We assumed effectiveness varied with the time of care-seeking but was constant across age and risk stratifications.

**Table 1.**
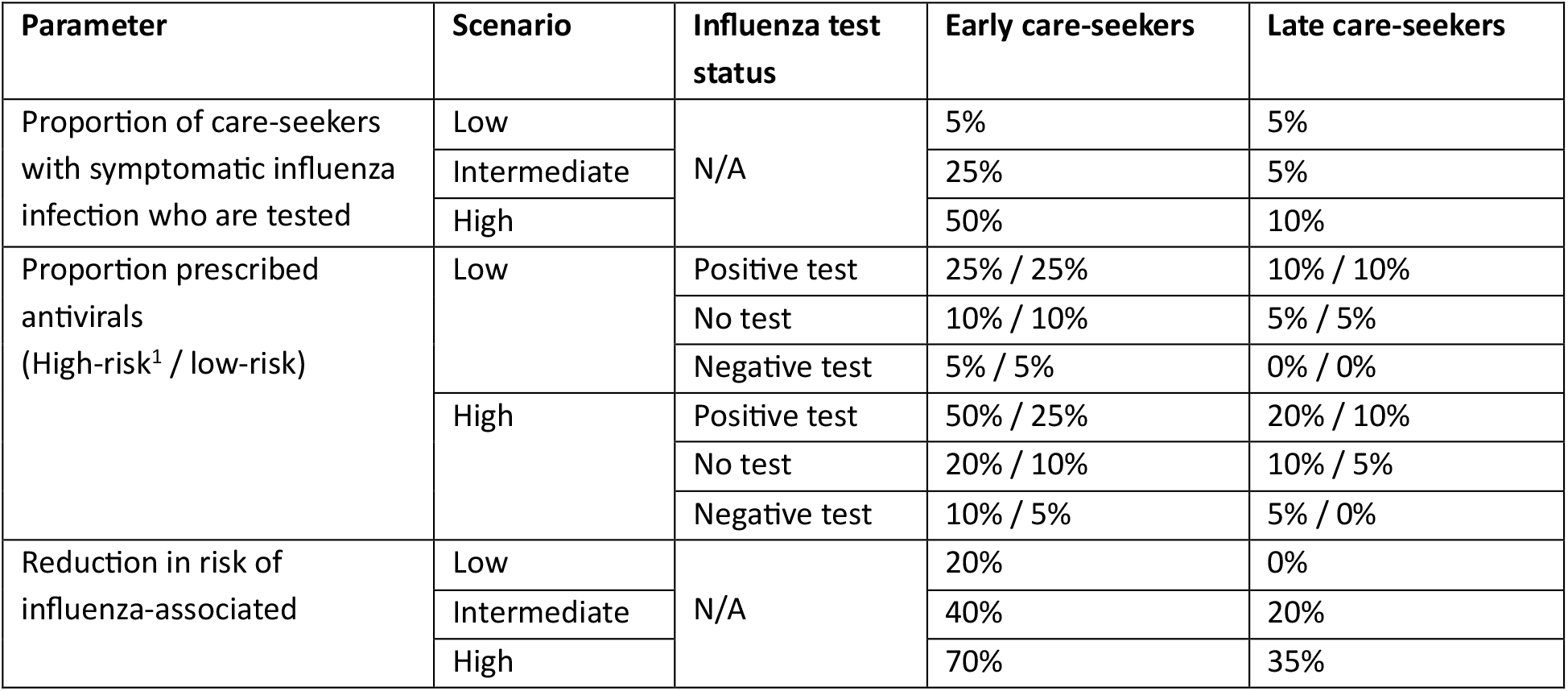
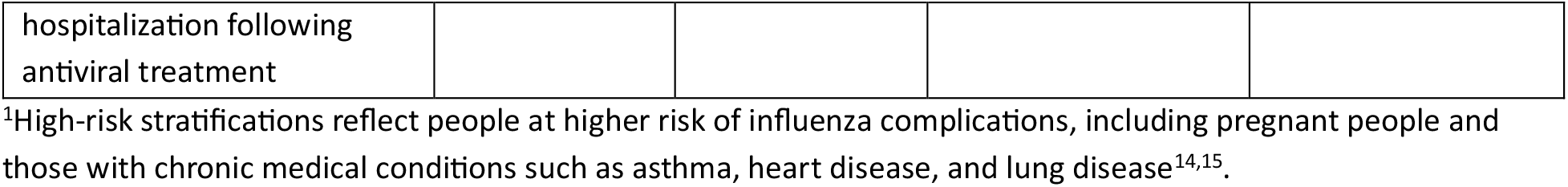
Initial scenario inputs for antiviral effectiveness and clinical testing and antiviral prescribing practices. Influenza antiviral effectiveness and testing values are assumed equal among low- and high-risk stratifications. All input values are assumed equal across age groups but may differ among early and late care-seekers.

| Parameter | Scenario | Influenza test status | Early care-seekers | Late care-seekers |
| --- | --- | --- | --- | --- |
| Proportion of care-seekers with symptomatic influenza infection who are tested | Low | N/A | 5% | 5% |
|  | Intermediate |  | 25% | 5% |
|  | High |  | 50% | 10% |
| Proportion prescribed antivirals (High-risk<sup>1</sup> / low-risk) | Low | Positive test | 25% / 25% | 10% / 10% |
|  |  | No test | 10% / 10% | 5% / 5% |
|  |  | Negative test | 5% / 5% | 0% / 0% |
|  | High | Positive test | 50% / 25% | 20% / 10% |
|  |  | No test | 20% / 10% | 10% / 5% |
|  |  | Negative test | 10% / 5% | 5% / 0% |
| Reduction in risk of influenza-associated | Low | N/A | 20% | 0% |
|  | Intermediate |  | 40% | 20% |
|  | High |  | 70% | 35% |
| hospitalization following<br>antiviral treatment |  |  |  |  |

<sup>1</sup>High-risk stratifications reflect people at higher risk of influenza complications, including pregnant people and those with chronic medical conditions such as asthma, heart disease, and lung disease<sup>14,15</sup>.

### Model simulation

Taking all combinations of possible values for testing, prescribing, and antiviral effectiveness resulted in 18 (3 × 2 × 3) initial scenarios (Table 1). All remaining model inputs were assumed to follow independent Uniform probability distributions, except population size and hospitalization risk which were defined by age and risk-stratified point estimates (Table S1). For each scenario, we generated 1,000 independent samples for the parameters with Uniform probability distributions (Figure 2). We then simulated the model for each of the 18,000 resulting combinations and calculated the number of influenza-associated hospitalizations that occurred among all symptomatic individuals. We also compared the estimated number of hospitalizations to a corresponding baseline scenario without antiviral medications. Results are presented as the number, or percent, of all hospitalizations averted relative to this baseline scenario. Finally, we tracked the number of antiviral courses prescribed in each simulation and calculated the number of prescriptions needed to avert one hospitalization as the total number of prescriptions divided by the total number of hospitalizations averted.

**Figure 2.**
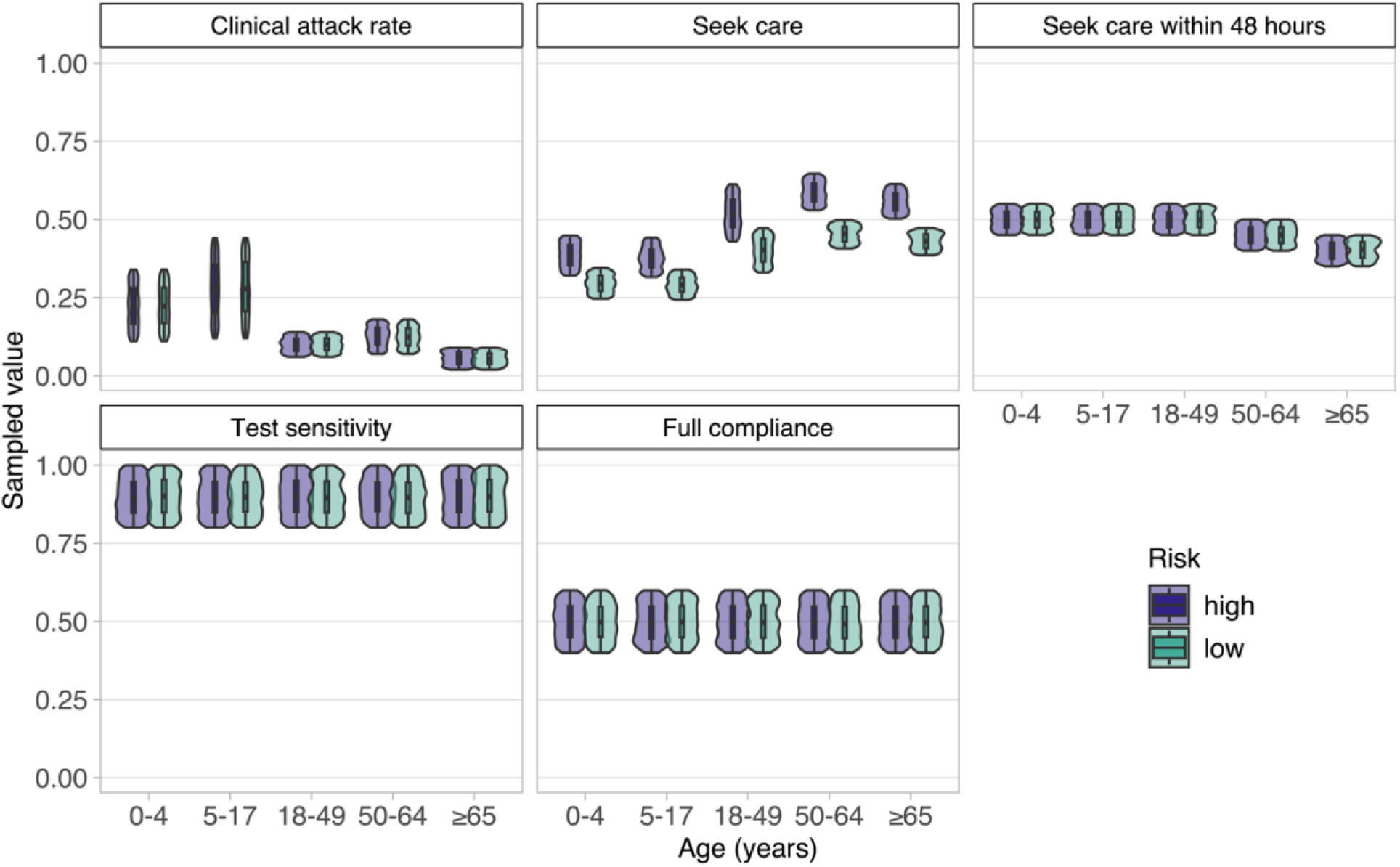
Simulated parameter inputs. Input parameter distributions were generated by taking 1,000 samples from their corresponding probability distributions. Risk stratifications refer to groups with high or low risk of influenza-associated hospitalization; full compliance refers to individuals who filled and completed a prescribed course of antiviral treatment.

### Alternative scenarios

In addition to the scenarios defined above, we considered alternative scenarios for improved uptake of influenza antivirals through increasing upstream levels of care-seeking, early care-seeking, testing, prescribing for those with a positive test, and the filling and completion of antiviral prescriptions (Figure 1). For each parameter and age group or risk stratification, we increased the initial scenario or sampled distributions by 50% (Figure S1, Table S2), and investigated the impact of increasing one parameter at a time or increasing multiple parameters simultaneously. For all alternative scenarios, we used the most optimistic initial levels of antiviral effectiveness, testing, and prescribing as the starting point (i.e., the initial scenario with the highest levels of effectiveness, testing, and prescribing).

All analyses were performed in R 4.0.3 using the `data.table` and `tidyverse` packages^30,31,32^.

## Results

### Initial scenarios

We estimated there would be an average of 531,920 influenza-associated hospitalizations (95^th^ percentile: 360,353–699,923) in the baseline scenario without influenza antivirals. The use of antivirals averted an average of 1,215 (802–1,693) hospitalizations in the least optimistic scenario with low testing, low prescribing, and low antiviral effectiveness, and 14,184 (8,700–20,224) hospitalizations in the most optimistic scenario with high testing, high prescribing, and high antiviral effectiveness (Figure S2A). These numbers corresponded to a modest percentage reduction in total hospitalizations: 0.2% (0.2– 0.3%) and 2.7% (2.2–3.2%) in the least optimistic and most optimistic scenarios, respectively. The total number of prescribed antiviral courses ranged from 1,370,274 (1,073,330–1,700,729) to 2,379,603 (1,914,821–2,879,333) and the number of prescriptions needed to avert one hospitalization ranged from 176 (117–261) to 1,168 (785–1,689) (Figure S2B).

Among individuals at higher risk of influenza complications, the number of averted hospitalizations increased with increasing age group, from an average of 45 (21–75) among children 0–4 years to 8,061 (2,927–14,216) among adults ≥65 years in the most optimistic scenario with high testing, high prescribing, and high antiviral effectiveness (Figure 3). When testing and prescribing were instead fixed at their lowest levels, the number averted ranged from 15 (7–26) among children 0–4 years to 2,849 (1,044–4,996) among adults ≥65 years. Notably, the percent of hospitalizations averted was greatest for adults aged 18–49 and 50–64 years, with the latter experiencing an average reduction of 3.7% (2.7–4.7%) in the most optimistic scenario (Figure S3). In all scenarios, the number of prescriptions needed to avert one hospitalization was highest among children 5–17 years and lowest among adults ≥65 years (Figure S4).

**Figure 3.**
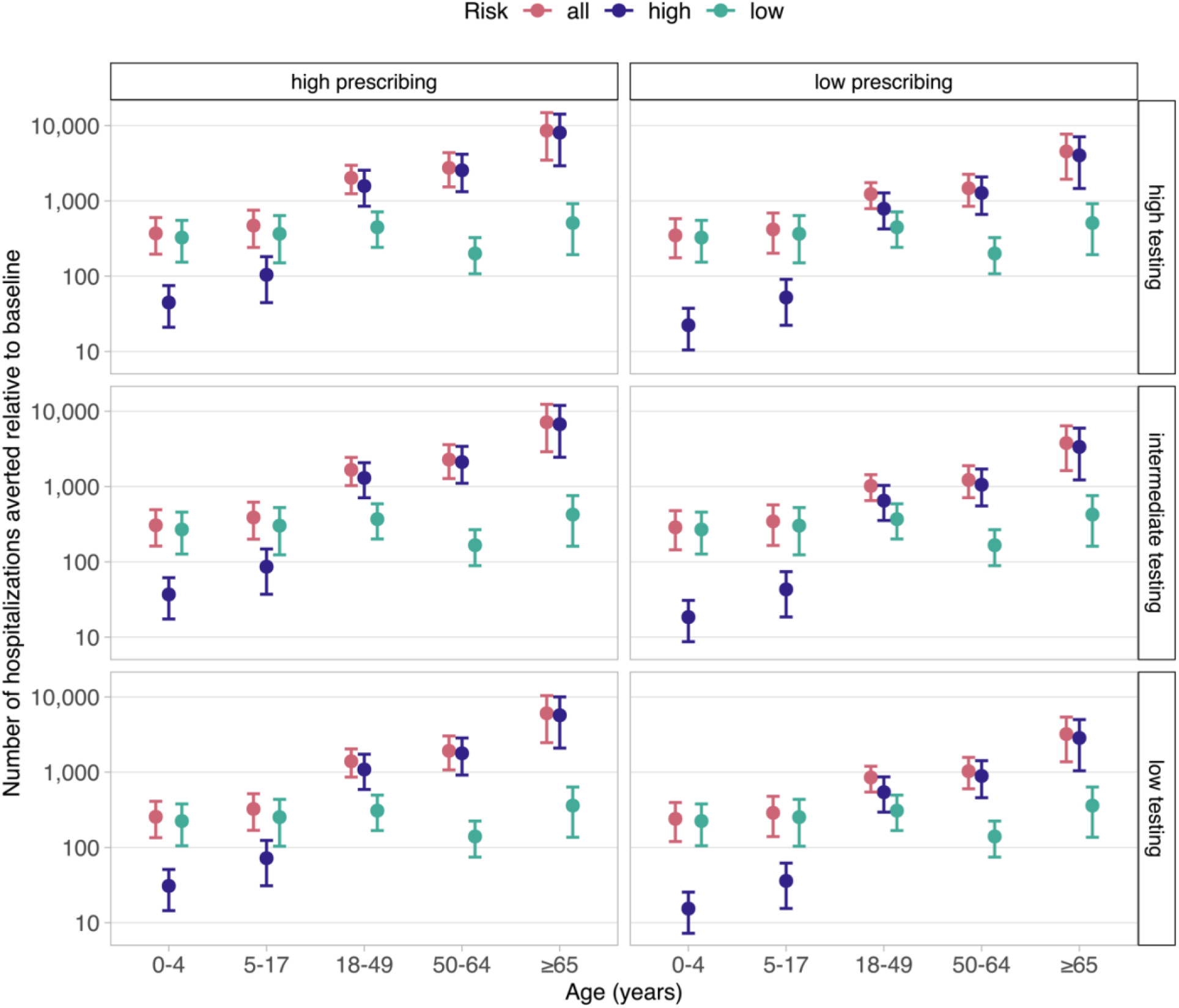
Estimated number of influenza-associated hospitalizations averted in each initial testing and prescribing scenario compared with a baseline scenario without antivirals. Antiviral effectiveness is fixed at its highest value (70% reduction in risk of hospitalization for early care-seekers and 35% for late care-seekers). Results are partitioned by age group and risk stratification, with points and error bars showing the mean and 95^th^ percentiles, respectively.

### Scenarios to improve antiviral impact

Given the relatively modest estimated impact of antivirals in reducing influenza-associated hospitalizations in the scenarios above, we explored whether this impact could be improved by increasing upstream levels of care-seeking, timing of care-seeking, testing, prescribing, or antiviral compliance. First, we considered the parameters with sampled distributions that were broadly associated with individual behavior and access to care and treatment (care-seeking, timing of care-seeking, and antiviral compliance) (Figure 1). Increasing either care-seeking or antiviral compliance by 50% resulted in equal improvements in averted hospitalizations compared to the most optimistic initial scenario. While increasing early care-seeking by 50% also resulted in improvements compared to the most optimistic initial scenario, these were slightly less than those achieved by increasing care-seeking or antiviral compliance (Figure 4). Unsurprisingly, the largest improvement was achieved by increasing all three parameters simultaneously. In this case, the number of averted hospitalizations was greatest among adults ≥65 years in the high-risk stratification, reaching up to 24,220 (8,709–42,968) (Figure 4A). In contrast, the greatest percent of hospitalizations were averted among adults aged 50–64 years in the high-risk stratification, reaching up to 11.1% (8.2–14.3%) (Figure 4B). The number of antiviral prescriptions needed to avert one hospitalization decreased in all alternative scenarios compared to the initial scenario, except when only care-seeking was improved (Figure 4C). The scenarios which resulted in the greatest decrease were those with increased antiviral compliance and with all parameters increased simultaneously.

**Figure 4.**
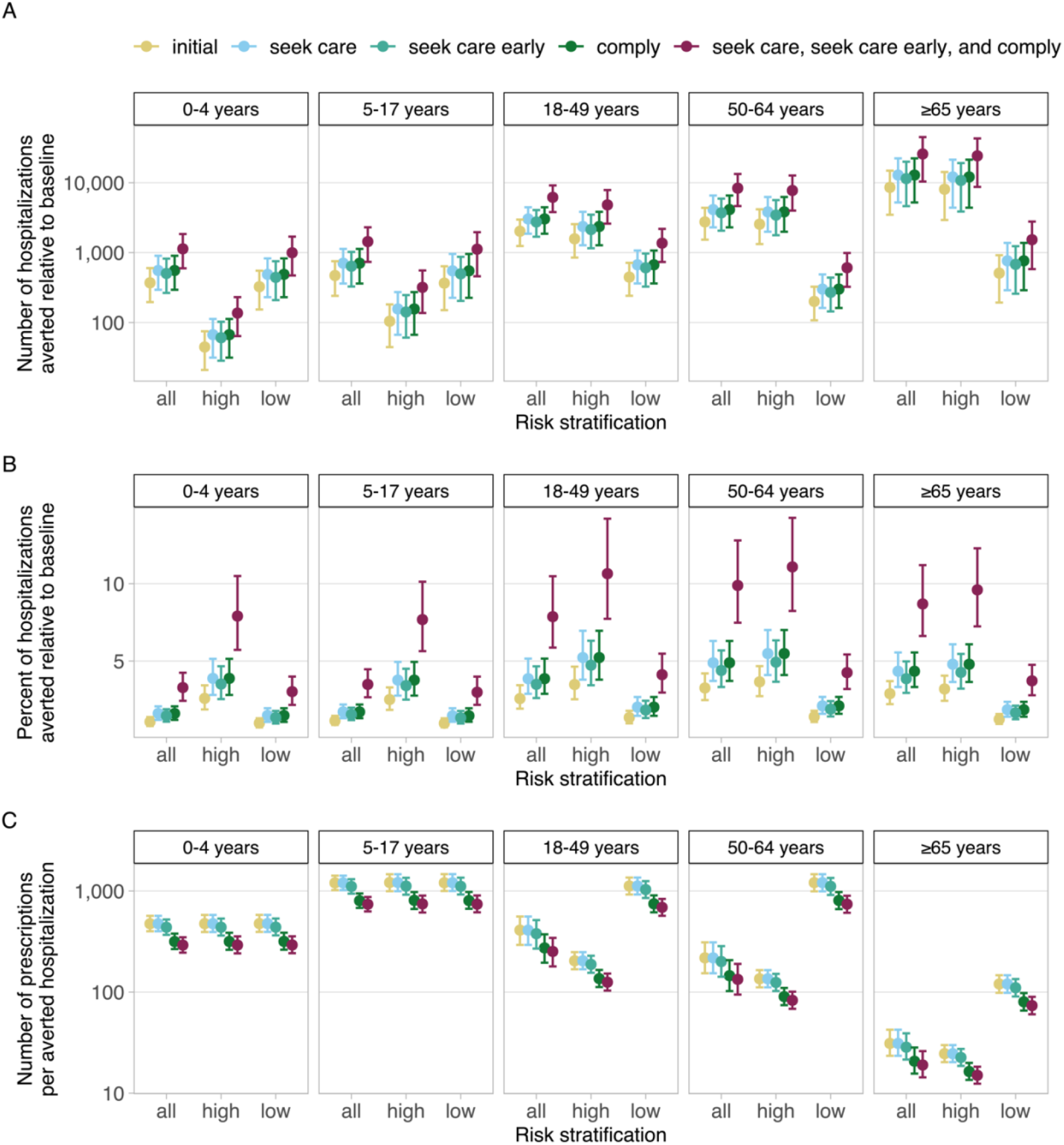
Estimated influenza-associated hospitalizations averted in select alternative scenarios compared with a baseline scenario without antivirals. (A) Number of hospitalizations averted. (B) Percent of hospitalizations averted. (C) Number of prescriptions needed to avert one hospitalization. Scenarios shown are as follows: most optimistic initial scenario (‘initial’); 50% increase in the fraction of people seeking care (‘seek care’); 50% increase in the fraction of people seeking care within 48 hours (‘seek care early’)’; 50% increase in the fraction of people completing an antiviral treatment course (‘comply’); and 50% increase in the fraction of people seeking care, seeking care early, and completing a treatment course. Antiviral effectiveness, testing, and prescribing are fixed at their highest initial values. Results are partitioned by age group and risk stratification, with points and error bars showing the mean and 95^th^ percentiles, respectively.

We also considered alternative scenarios for the parameters broadly associated with clinical practices, i.e., the probability of testing and prescribing (Figure 1). Increased antiviral prescribing among outpatients with a positive test averted more hospitalizations than increased testing and required slightly fewer prescriptions to avert one hospitalization (Figure S5). Increasing both parameters averted the most hospitalizations among adults ≥65 years in the high-risk stratification (up to 12,909 [4,650– 22,834]), and the greatest percent of hospitalizations among adults 50–64 years in the high-risk stratification (5.9% [4.4–7.6%]).

Finally, to explore even greater potential improvements in antiviral impact, we considered an alternative scenario in which all five parameters were increased simultaneously. Again, the number of averted hospitalizations was greatest among adults ≥65 years in the high-risk stratification, at 40,093 (14,332–71,087), whereas the greatest percentage of hospitalizations was averted among adults 50–64 years in the high-risk stratification, at 18.5% (13.5–24.0%) (Figure S6). Across all age and risk groups, the use of antivirals averted 71,142 (43,581–102,969) hospitalizations, or 13.3% (11.1–16.2%), and the overall number of prescriptions needed to avert one hospitalization was 107 (71–157).

## Discussion

We developed a probabilistic model to estimate the impact of antivirals in reducing hospitalizations among outpatients with symptomatic influenza virus infection under a range of plausible parameter values. Antiviral usage reduced hospitalizations compared to a scenario without antivirals. Although the percentage reduction (0.2–2.7%) was modest, the absolute reduction (1,215–14,184) could be clinically significant and supports the use of antivirals to treat outpatients with suspected or confirmed influenza. Among age groups and risk stratifications, the greatest impacts occurred among adults ≥65 years and individuals with conditions associated with higher risk of influenza complications. Our results align with CDC recommendations to prioritize these individuals for antiviral treatment. More broadly, our framework can help identify groups for which treatment could have the greatest impact, particularly in situations where antiviral resources may be limited.

In all our scenarios we assumed a non-zero benefit of antivirals in reducing influenza-associated hospitalizations among early care-seekers who completed a full treatment course: from 20% in the least optimistic scenario to 70% in the most optimistic scenario. The variation in these values reflects underlying challenges in estimating the true impact of antivirals on preventing influenza-associated hospitalizations, with randomized controlled trials (RCTs) requiring very large sample sizes (e.g., tens of thousands of patients) for sufficient statistical power. For example, a recent meta-analysis of RCTs for oseltamivir that reported no significant reduction in hospitalizations among outpatients with influenza was likely underpowered and, in addition, included trials in which hospitalization was not the primary endpoint^29,33,34^. Given these challenges, observational studies and RCTs that investigate alternative end points for disease progression can provide valuable additional evidence to support treatment guidelines. Examples include a meta-analysis of observational data that demonstrated a significant reduction in the odds of hospital admission among adults and children treated with neuraminidase inhibitors (including oseltamivir) during the 2009 H1N1 pandemic^3^, and another meta-analysis of RCTs that found oseltamivir significantly reduced the risk of lower respiratory tract complications requiring antibiotics among adults treated within 36 hours of illness onset^2^. These studies suggest that antivirals, like oseltamivir, can reduce the risk of severe disease progression when treatment is initiated early after influenza symptom onset. We therefore focused on a positive effect of antivirals in reducing hospitalization risk but did include a zero benefit among late care-seekers (in the least optimistic scenario) to incorporate the possibility of no effect when treatment is delayed. Our scenarios were also conservative in assuming no benefit of antivirals among outpatients who did not complete a full treatment course. Thus, our results show a positive impact of antivirals in averting hospitalizations across a range of plausible antiviral effect magnitudes, from conservative to optimistic, and support their use in outpatient settings to reduce severe influenza disease burden, particularly among patients at higher risk of complications.

As antiviral effectiveness was just one parameter influencing averted influenza-associated hospitalizations, we also explored the effect of improving five other parameters (including care-seeking, testing, and prescribing) that determined whether someone received, and ultimately completed, an antiviral prescription. Over 71,000 (13.3%) hospitalizations were averted when all parameters were increased by 50%, and the number of prescriptions needed to avert one hospitalization was reduced. Thus, combined improvements in multiple components of the care and treatment pathway could substantially improve the impact of antivirals in reducing influenza-associated hospitalizations.

Our framework has several limitations. First, we modeled the direct effects of antivirals in reducing the risk of hospitalization among treated outpatients but did not account for indirect effects that may occur following reductions in the risk of onward influenza transmission. However, these effects may be small as prior work has shown oseltamivir treatment does not significantly reduce influenza viral shedding^35,36^. In line with this, a mathematical modeling study that did account for indirect effects still found modest impacts of antivirals in reducing influenza-associated hospitalizations^37^. Second, we did not consider the prophylactic use of antivirals to reduce infection risk in individuals exposed to an influenza virus. Such measures may be more important in congregate settings, like long-term care facilities^38^, which were not the focus of this work. Third, our estimates of the proportion of the population at higher risk of influenza complications did not include people with a BMI ≥40 or those living in long-term care facilities^5,15^. Accounting for these groups would expand the population at increased risk for influenza complications, among whom antivirals had the greatest impact, and thus increase the estimated number of averted hospitalizations. Fourth, our test sensitivity inputs were based on molecular tests (rapid or standard) rather than rapid antigen tests which typically have lower sensitivity (e.g., 50–70%)^39^. Allowing a fraction of tests to be rapid antigen would reduce the number of tested individuals who receive a positive result, and thus reduce the number of antiviral medications ultimately prescribed. However, the magnitude of the reduction would depend on the relative proportions of antigen and molecular tests, which are highly variable in outpatient settings^21^. Fifth, we assumed that the risk of influenza-associated hospitalization (in the absence of antiviral treatment) was the same among people who did and did not seek care. This risk was parameterized using case-hospitalization ratios that are adjusted for care-seeking behavior and thus reflect an average across all individuals with influenza-associated illness. More generally, our model inputs will likely vary by many factors, including location, clinical setting, type of antiviral medication, and access to insurance and medical care. We included probabilistic ranges or different scenarios to account for this variation, but our results should not be interpreted as an estimate of the current impact of antivirals on influenza-associated hospitalizations. Instead, they provide a range of plausible estimates given likely underlying heterogeneity in individual care-seeking, clinical testing, and antiviral prescribing practices across the United States.

We modeled the potential impact of antiviral treatment in reducing hospitalizations among outpatients with influenza. In general, we estimated modest overall reductions in hospitalizations, but found greater benefits among people at higher risk of influenza-associated complications, and when multiple components of the care and treatment pathway were improved. Our findings align with current guidance for prioritizing prompt antiviral treatment among persons with suspected or confirmed influenza who are at higher risk of influenza complications and, more generally, demonstrate the utility of our framework in assessing the potential impact of influenza antivirals in a large population with uncertain parameter inputs.

## Data Availability

All model inputs and code needed to perform the analysis will be made available at https://github.com/CDCgov upon publication. Data were used solely to inform model inputs and were the result of secondary analyses; the original sources are cited in the text.

## Acknowledgements

The authors wish to thank Matthew Gilmer, Ryan Threlkel, Daniel Moore, and Alexandra Mellis for data contributions that informed model inputs and Pragati Prasad for early contributions to the model code.

## Conflict of Interest Statement

The authors have no conflicts of interest or funding sources to declare.

## Notes

### Competing Interest Statement

The authors have declared no competing interest.

### Funding Statement

This study did not receive any external funding.

