## Supplementary Information for "Modeling the potential impacts of outpatient antiviral treatment in reducing influenza-associated hospitalizations in the United States"

**Table S1. Initial inputs for parameters with fixed values or probabilistic distributions.** Ranges represent the lower and upper limits of a Uniform probability distribution.

| Parameter | Age (years) | Input value or range | Ratio between high-risk: low-risk stratifications | Source(s) |
|---|---|---|---|---|
| Population | 0–4 | 22,221,466 | N/A | [1] |
| | 5–17 | 53,912,474 | | |
| | 18–49 | 140,148,894 | | |
| | 50–64 | 62,892,984 | | |
| | ≥65 | 57,794,852 | | |
| Proportion at higher risk of influenza complications | 0–4 | 0.05 | N/A | [2] |
| | 5–17 | 0.10 | | |
| | 18–49 | 0.20 | | |
| | 50–64 | 0.35 | | |
| | ≥65 | 0.55 | | |
| Risk of influenza-associated hospitalization | 0–4 | 0.007 | 1 | [3,4] |
| | 5–17 | 0.003 | 1 | |
| | 18–49 | 0.006 | 5.5 | |
| | 50–64 | 0.011 | 8.9 | |
| | ≥65 | 0.091 | 4.9 | |
| Influenza clinical attack rate | 0–4 | 0.11–0.34 | 1 (no difference) | [5] |
| | 5–17 | 0.12–0.44 | | |
| | 18–49 | 0.06–0.14 | | |
| | 50–64 | 0.07–0.18 | | |
| | ≥65 | 0.02–0.09 | | |
| Proportion seeking care | 0–4 | 0.25–0.35 | 1.3 for all age groups | [6,7] |
| | 5–17 | 0.25–0.35 | | |
| | 18–49 | 0.35–0.50 | | |
| | 50–64 | 0.45–0.55 | | |
| | ≥65 | 0.45–0.55 | | |
| Proportion seeking care within 48 hours | 0–4 | 0.45–0.55 | 1 (no difference) | [6–8] |
| | 5–17 | 0.45–0.55 | | |
| | 18–49 | 0.45–0.55 | | |
| | 50–64 | 0.40–0.50 | | |
| | ≥65 | 0.35–0.45 | | |
| Influenza test sensitivity | N/A | 0.80–1.00 | 1 (no difference) | [9,10] |
| Proportion filling and completing an antiviral prescription course | N/A | 0.40–0.60 | 1 (no difference) | [7,11] |

**Table S2. Alternative scenario inputs for testing and prescribing probabilities.**

| Parameter | Risk stratification | Time of care-seeking | Original value | Alternative value |
|---|---|---|---|---|
| Proportion tested | N/A | Early | 50% | 75% |
| | | Late | 10% | 15% |
| Proportion with a positive influenza test who are prescribed antivirals | High-risk | Early | 50% | 75% |
| | | Late | 20% | 30% |
| | Low-risk | Early | 25% | 37% |
| | | Late | 10% | 15% |

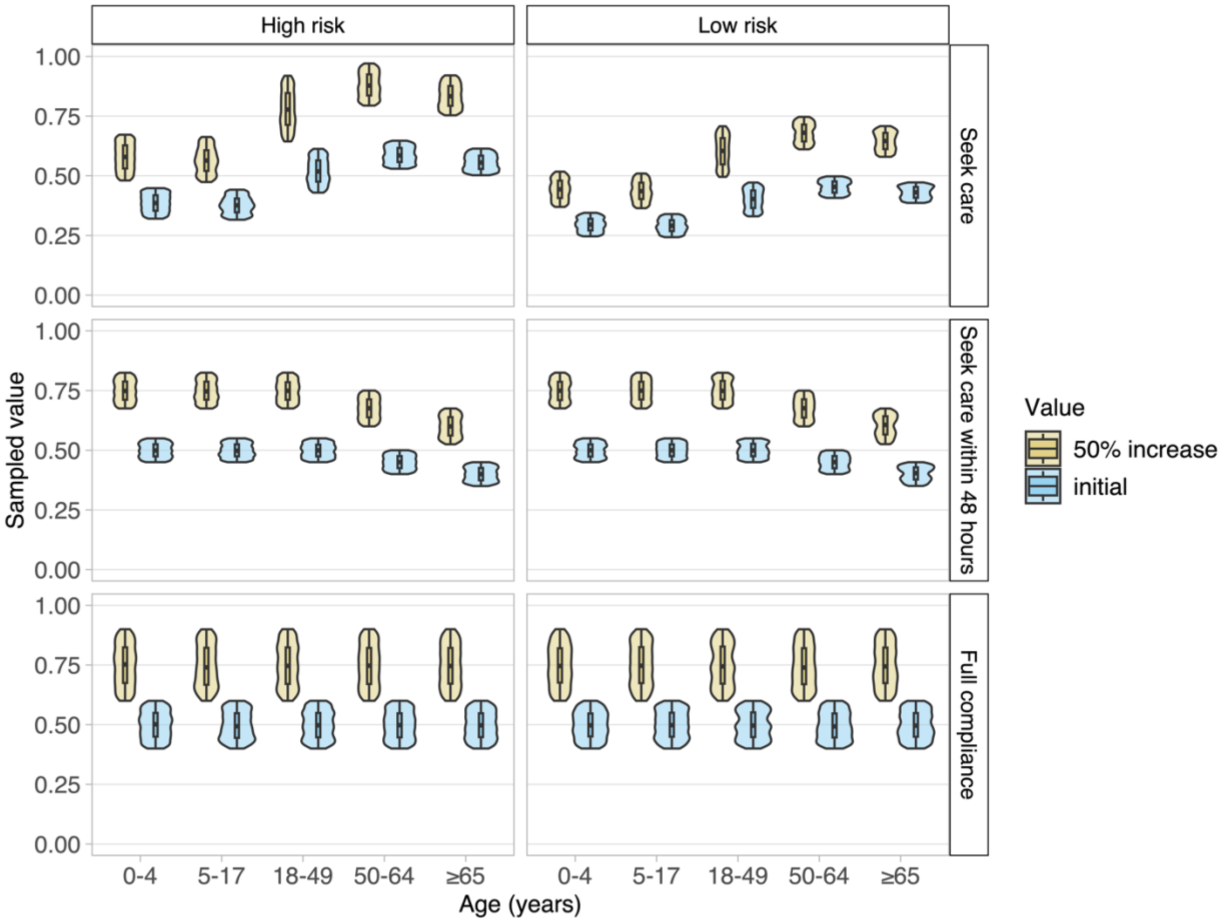


**Figure S1. Alternative scenario inputs for parameters with sampled distributions: care-seeking, time of care-seeking, and antiviral compliance.** The alternative scenarios represent a 50% increase in all values sampled for each parameter.

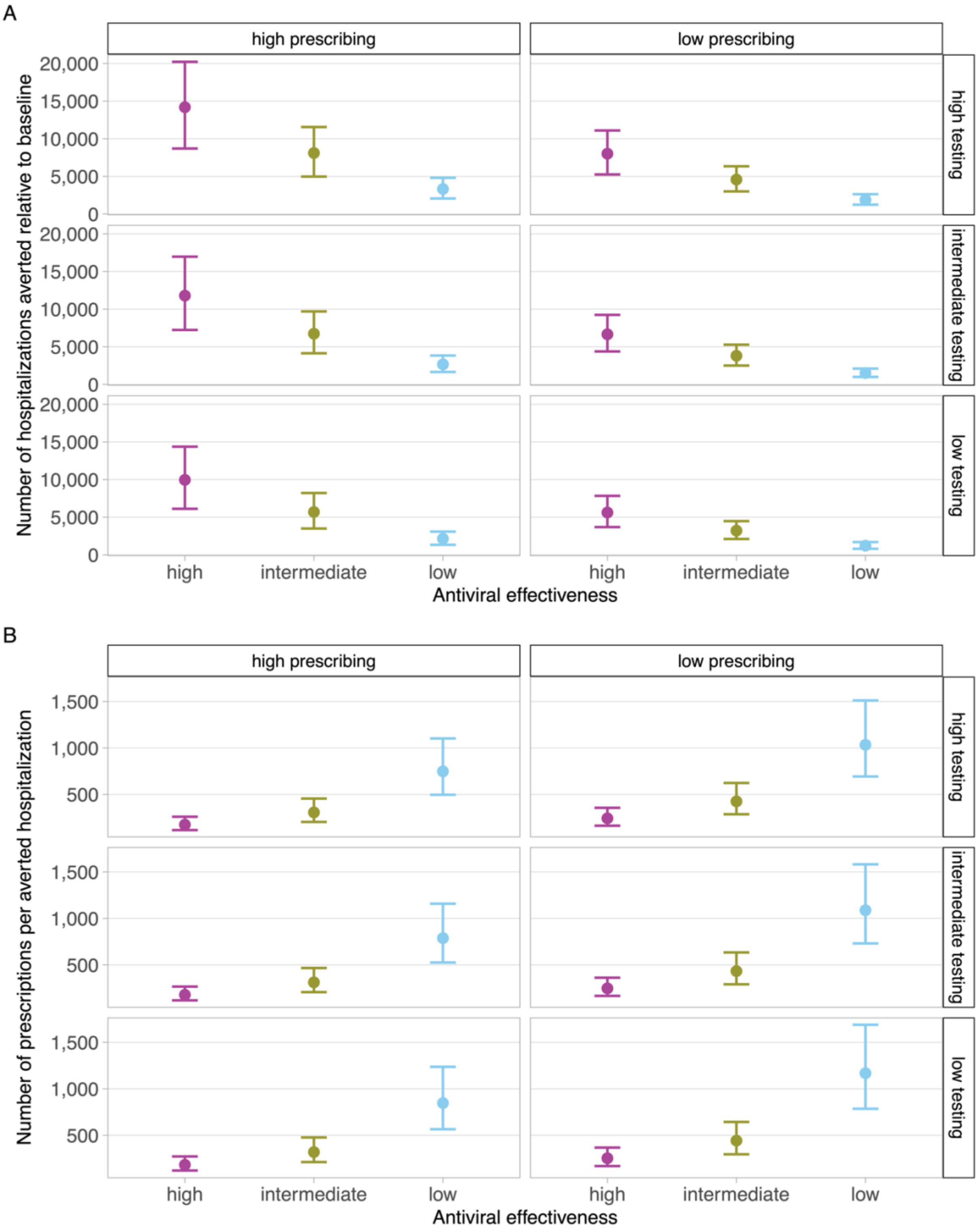


**Figure S2. Estimated number of influenza-associated hospitalizations averted and antiviral prescriptions needed in each initial scenario compared with a baseline scenario without antivirals.** (A) Total number of hospitalizations averted. (B) Number of antiviral prescriptions needed to avert one hospitalization. Results are combined across age groups and risk stratifications. Points and error bars show the mean and 95th percentiles, respectively.

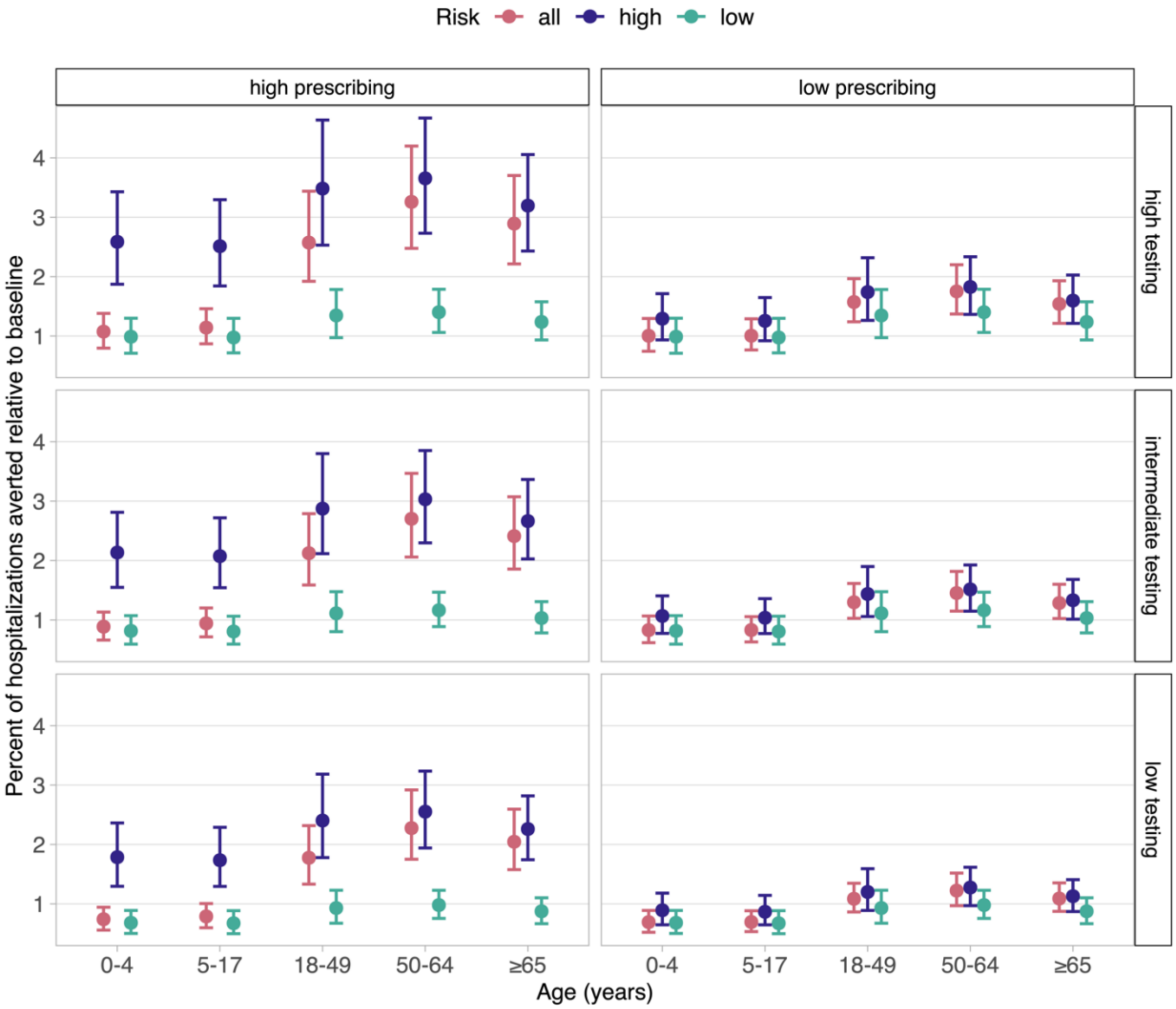


**Figure S3. Estimated percent of influenza-associated hospitalizations averted in each initial scenario compared with a baseline scenario without antivirals.** Antiviral effectiveness is fixed at its highest value (70% reduction in risk of hospitalization for early care-seekers and 35% for late care-seekers). Results are partitioned by age group and risk stratification. Points and error bars show the mean and $95^{th}$ percentiles, respectively.

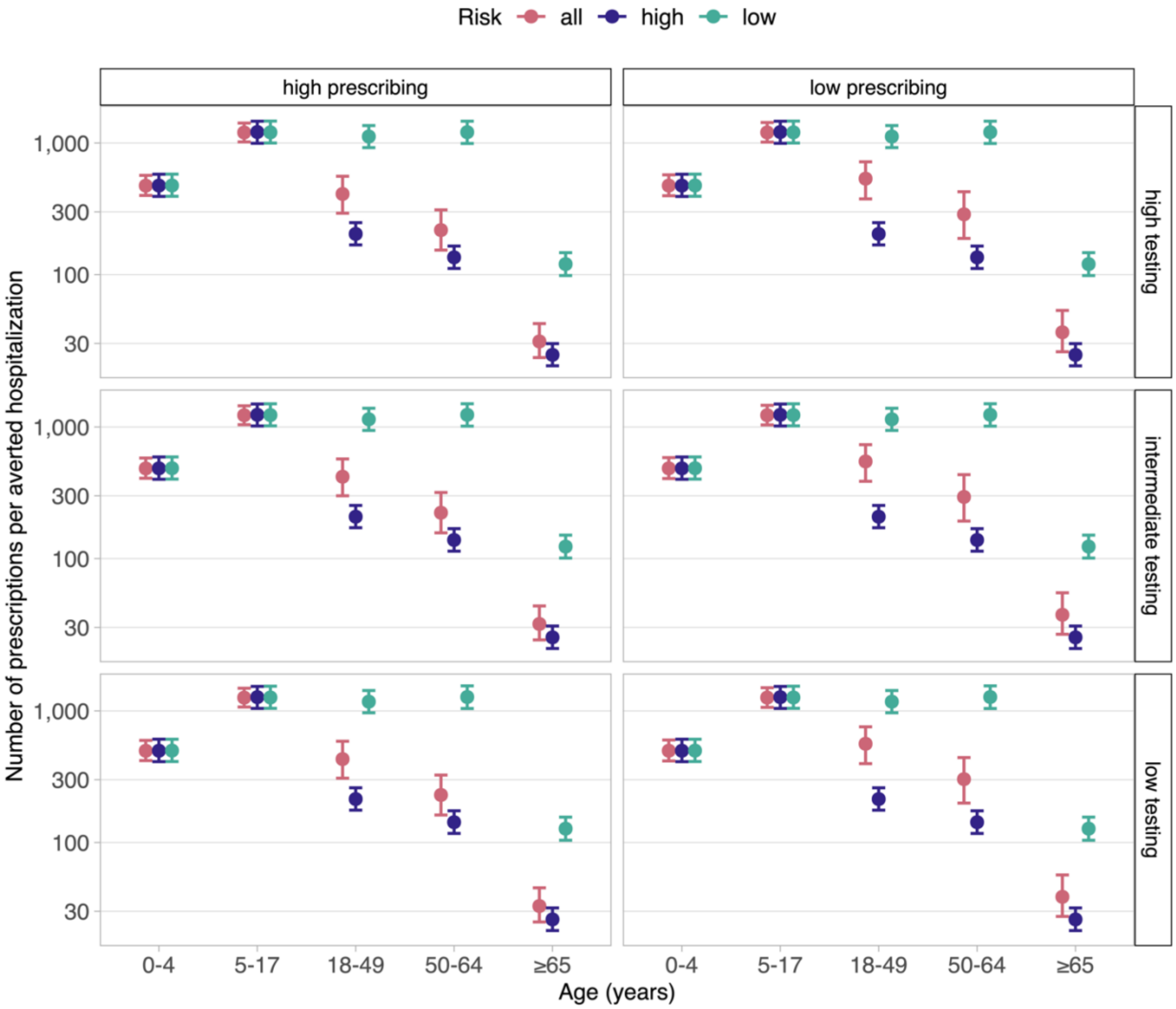


**Figure S4. Estimated number of antiviral prescriptions needed to avert one influenza-associated hospitalization.** Antiviral effectiveness is fixed at its highest value (70% reduction in risk of hospitalization for early care-seekers and 35% for late care-seekers). Results are partitioned by age group and risk stratification. Points and error bars show the mean and 95$^{th}$ percentiles, respectively.

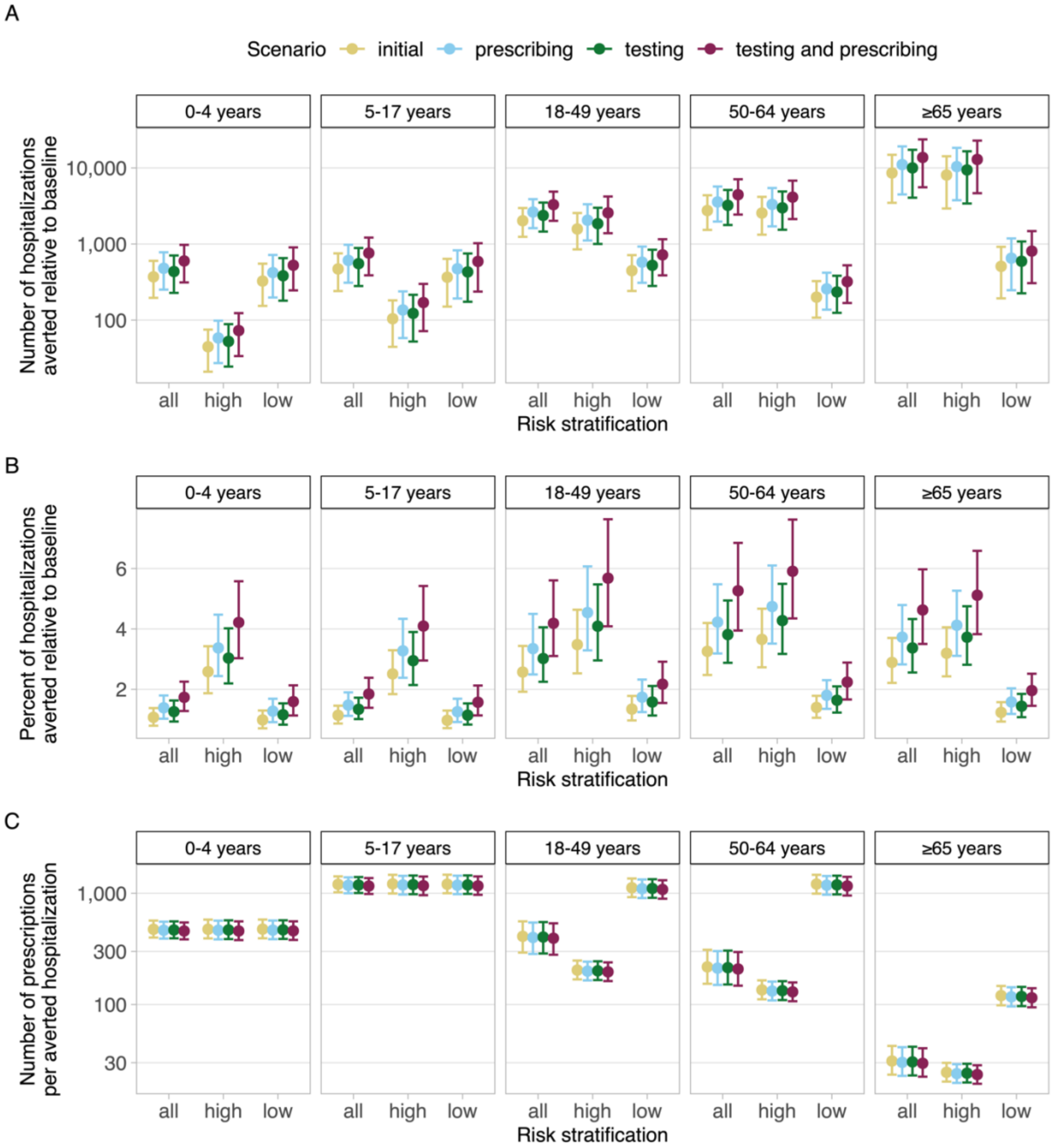


**Figure S5. Estimated impact of antivirals in select alternative scenarios compared with a baseline scenario without antivirals.** (A) Number of hospitalizations averted. (B) Percent of hospitalizations averted. (C) Number of prescriptions needed to avert one hospitalization. Scenarios are as follows: the most optimistic initial scenario ('initial'); a 50% increase the probability of testing ('testing'); a 50% increase in the probability of prescribing among people with a positive test ('prescribing'); and a 50% increase in both the probability of testing and the probability of prescribing among people with a positive test ('testing and prescribing'). In all scenarios, antiviral effectiveness is fixed at its highest value (70% reduction in risk of hospitalization for early care-seekers and 35% for late care-seekers). Results are partitioned by age group and risk stratification. Points and error bars show the mean and 95$^{th}$ percentiles, respectively. Hospitalizations refer to influenza-associated hospitalizations.

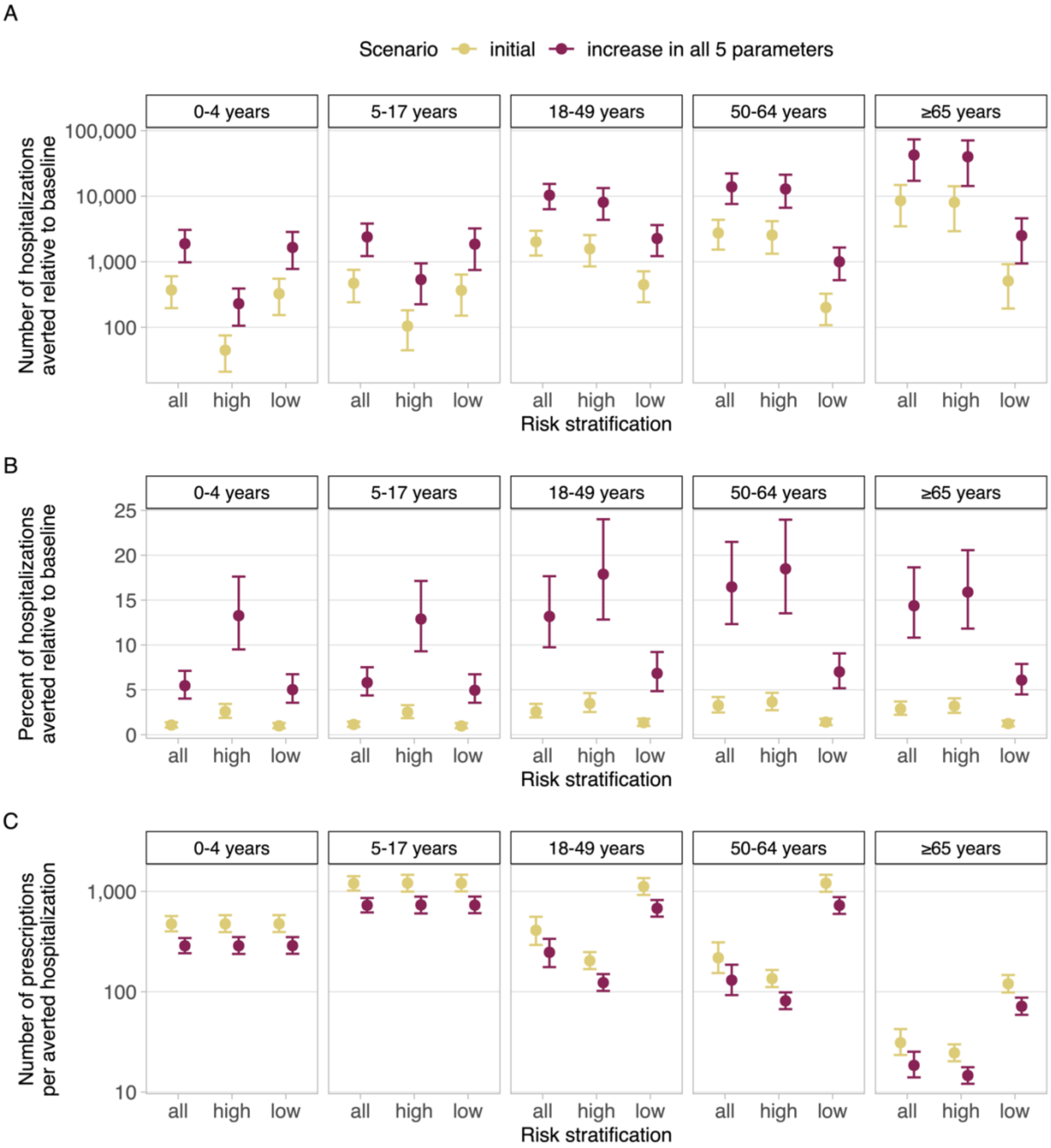


**Figure S6. Estimated impact of antivirals in an initial and alternative scenario compared with a baseline scenario without antivirals.** (A) Number of hospitalizations averted. (B) Percent of hospitalizations averted. (C) Number of prescriptions needed to avert one hospitalization. Depicted scenarios are: the most optimistic initial scenario ('initial') and a scenario with a 50% increase in five model parameters — the fraction of people seeking care, the fraction of people seeking care within 48 hours, the fraction of people completing an antiviral treatment course, the probability of testing, and the probability of prescribing among people with a positive test. In all scenarios, antiviral effectiveness is fixed at its highest value (70% reduction in risk of hospitalization for early care-seekers and 35% for late care-seekers). Results are partitioned by age group and risk stratification. Points and error bars show the mean and 95$^{th}$ percentiles, respectively. Hospitalizations refer to influenza-associated hospitalizations.

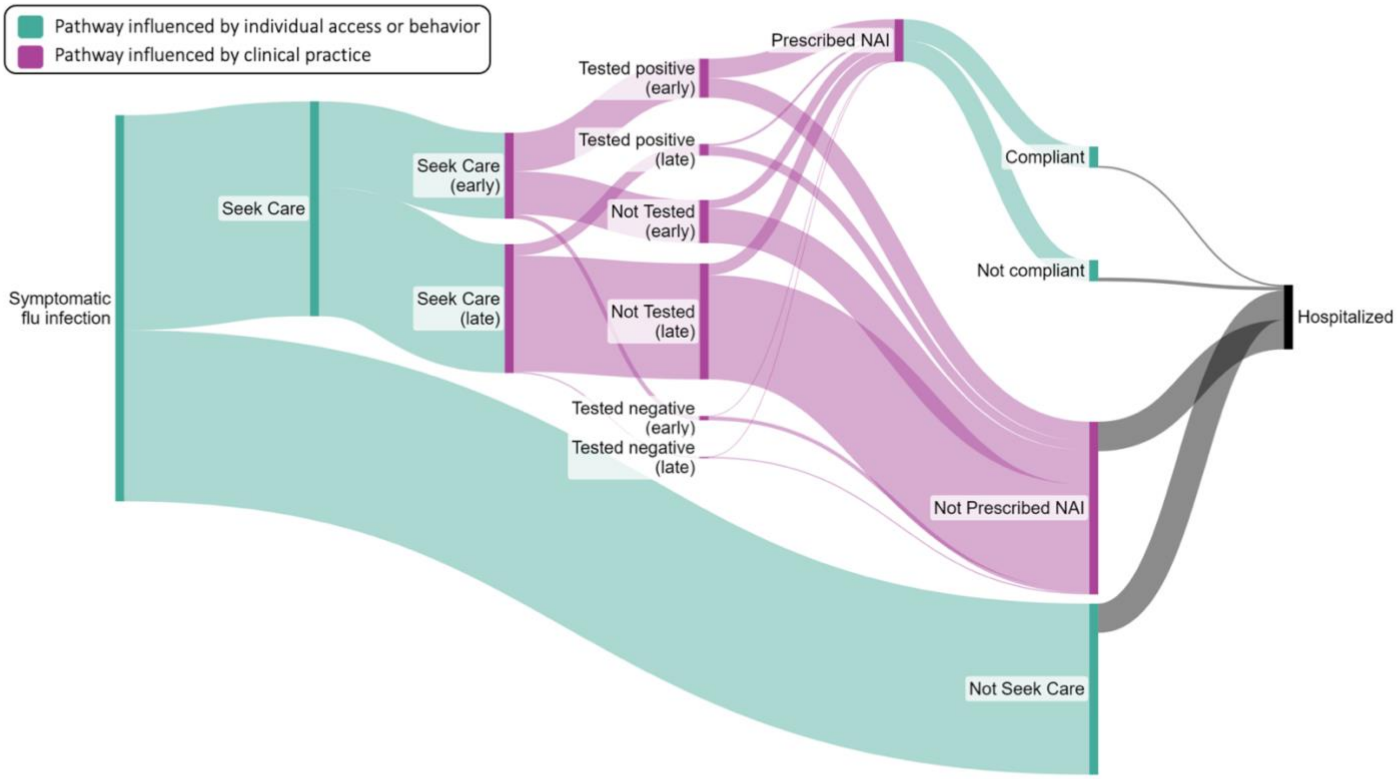


**Figure S7. Sankey diagram showing example numbers of individuals progressing through each pathway. From symptomatic influenza ('flu') infection to hospitalization.** Example values were chosen for adults ≥65 years who are at higher risk of influenza complications. Note that antiviral effectiveness varies by the timing of care-seeking and will lead to different numbers progressing through the fully compliant to hospitalized pathway. The colors indicate pathways that are influenced by parameters relating to individual behavior and/or access to medical care and treatment (care-seeking, timing of care-seeking, and antiviral compliance) or parameters relating to clinical testing and prescribing practices. 'NAI' refers to a neuraminidase inhibitor antiviral such as oseltamivir. Figure created at sankeymatic.com[12].
